# The clinical spectrum of COVID-19: A population-based cohort study in Iceland

**DOI:** 10.1101/2020.08.09.20171249

**Authors:** Elias Eythorsson, Dadi Helgason, Ragnar Freyr Ingvarsson, Helgi K Bjornsson, Lovisa Bjork Olafsdottir, Valgerdur Bjarnadottir, Hrafnhildur Linnet Runolfsdottir, Solveig Bjarnadottir, Arnar Snaer Agustsson, Kristin Oskarsdottir, Hrafn Hliddal Thorvaldsson, Gudrun Kristjansdottir, Brynja Armannsdottir, Agnar Bjarnason, Birgir Johannsson, Olafur Gudlaugsson, Magnus Gottfredsson, Martin I Sigurdsson, Olafur S Indridason, Runolfur Palsson

## Abstract

**Background:** Previous studies on the epidemiology and clinical characteristics of COVID-19 have generally been limited to hospitalized patients. The aim of this study was to describe the complete clinical spectrum of COVID-19, based on a nationwide cohort with extensive diagnostic testing and a rigorous contact tracing approach.

**Methods:** A population-based cohort study examining symptom progression using prospectively recorded data on all individuals with a positive test (RT-PCR) for severe acute respiratory syndrome coronavirus 2 (SARS-CoV-2) who were enrolled in a telehealth monitoring service provided to all identified cases in Iceland. Symptoms were systematically monitored from diagnosis to recovery.

**Results:** From January 31 to April 30, 2020, a total of 45,105 individuals (12% of the Icelandic population) were tested for SARS-CoV-2, of whom 1797 were positive, yielding a population incidence of 5 per 1000 individuals. The most common presenting symptoms were myalgia (55%), headache (51%), and non-productive cough (49%). At the time of diagnosis, 5.3% of cases reported no symptoms and 3.1% remained asymptomatic during follow-up. In addition, 216 patients (13.8%) and 349 patients (22.3%) did not meet the case definition of the Centers for Disease Control and Prevention and the World Health Organization, respectively. The majority (67.5%) of patients had mild symptoms throughout the course of the disease.

**Conclusion:** In the setting of broad access to diagnostic testing, the majority of SARSCoV-2-positive patients were found to have mild symptoms. Fever and dyspnea were less common than previously reported. A substantial proportion of patients did not meet recommended case definitions at the time of diagnosis.

**Key points:** *Question:* What is the frequency and progression of various symptoms experienced by patients with COVID-19?

*Findings:* In this population-based cohort study that included all SARS-CoV-2-positive patients in Iceland, most patients (67.5%) had mild symptoms throughout their disease course. At the time of diagnosis, 5.3% were asymptomatic, of whom roughly half developed symptoms during follow-up. Common presenting symptoms included myalgia (55%), headache (51%), and non-productive cough (49%). At diagnosis, 13.8% and 22.3% did not meet the Centers for Disease Control and Prevention and World Health Organization case definitions for suspected COVID-19, respectively.

*Meaning:* In the setting of broad access to diagnostic testing, the majority of SARSCoV-2-positive patients were found to have mild symptoms and almost one-fifth did not meet published clinical criteria for RT-PCR testing.

## Introduction

On December 31 2019, the first cases of an atypical pneumonia of unidentified etiology were reported in Wuhan, China.^1^ One week later, a novel betacoronavirus, later named severe acute respiratory syndrome coronavirus (SARS-CoV-2), was identified as the causative pathogen^2,3^, and the disease subsequently termed coronavirus disease 2019 (COVID-19). The World Health Organization (WHO) declared the COVID-19 outbreak a pandemic on March 11, 2020.^4^

COVID-19 has a wide range of clinical manifestations, ranging from an asymptomatic state or mild respiratory symptoms to severe viral pneumonia and acute respiratory distress syndrome.^5-7^ Previous publications have suggested that approximately 81% of patients have mild symptoms, 14% have severe symptoms, and 5% become critically ill.^8^ Besides respiratory symptoms, dysosmia, dysgeusia, abdominal pain, diarrhea, and rash have been described.^5,9^ Most published studies on the clinical characteristics of COVID-19 have been retrospective^5,8,10,11^ and limited to inpatients^12,13^ and therefore do not capture the full clinical spectrum of the disease.

The first case of COVID-19 in Iceland was diagnosed on February 27, 2020.^14^ Icelandic health authorities responded immediately by isolating patients and instituting systematic contact tracing and quarantine of exposed individuals.^15^ Broad access to diagnostic testing became available in Iceland early in the course of the pandemic, allowing the highest rate of SARS-CoV-2 testing in the world.^16^ Approximately one month after the first case was identified, the incidence of undetected cases was found to be only 0.6% using random population screening.^14^ All SARS-CoV-2-positive individuals were actively monitored at a newly established COVID-19 outpatient clinic at Landspitali–The National University Hospital (LUH).^15,17^ The contact tracing and containment strategies implemented by the Icelandic authorities rapidly curbed the epidemic, with only 4 new cases diagnosed between May 1 and 15.^14^

In this paper, we describe the analysis of prospectively collected data on all SARS-CoV2-positive patients in Iceland and characterize the epidemiology and full clinical spectrum of COVID-19 in a nationwide cohort.

## Methods

### Study population and design

This population-based cohort study included all patients who tested positive for SARSCoV-2 between February 27 and April 30, 2020, and were actively monitored at LUH. All individuals in Iceland who tested positive for SARS-CoV-2 were immediately contacted, instructed to isolate, and enrolled in a telehealth monitoring service. Monitoring involved frequent telephone interviews by a nurse or physician, through which the patient’s clinical status was evaluated. From February 27 to March 16, 2020, the content and documentation of these interviews was at the discretion of the nurse or physician making the call. On March 17, a standardized data entry form was built directly into the national electronic medical record system, facilitating a structured approach to the clinical evaluation of COVID-19 patients. The study was approved by the National Bioethics Committee (VSN-20-078).

### Virological testing

Real-time reverse-transcriptase polymerase chain reaction (RT-PCR) was used for detection of SARS-CoV-2 RNA in nasopharyngeal and oropharyngeal swabs, based on the WHO recommended protocol from Charité, Berlin^18^ or using a commercial kit (TaqMan 2019-nCoV Assay from Thermo Fisher Scientific) as previously described.^15^ Three testing protocols were implemented: targeted testing, open-invitation population screening, and random population screening. Targeted testing was performed at the Department of Clinical Microbiology at LUH, whereas population screening was carried out by deCODE genetics, a biopharmaceutical company based in Reykjavik.^15^ Targeted testing began on January 31, 2020, and included clinically suspected cases and individuals at high risk of exposure. Open-invitation population screening began on March 13, 2020, and was available to all Icelandic residents who were not currently quarantined and did not have symptoms that would prompt targeted testing. Finally, a randomly chosen sample of 6782 Icelanders was offered testing via telephone text message on March 31 and April 1, 2020, of whom 2283 were included.

### Data collection

To confirm the completeness of telehealth enrollment, results of all SARS-CoV-2 testing were obtained from databases of LUH and deCODE genetics. Negative samples were used for denominator calculations. Baseline characteristics of SARS-CoV-2-positive patients and longitudinal data on symptom progression were obtained from the standardized data entry forms used by the COVID-19 Clinic. Data on clinical outcomes were extracted from LUH database. Data were linked using government issued national identification numbers. Population demographics were obtained from Statistics Iceland (www.statice.is).

### The telehealth monitoring service

The initial patient interviews were conducted by a physician who informed the patients of the diagnosis, evaluated their health, and instructed them to self-isolate at home. A checklist of 19 specific symptoms was presented to each patient during the initial and all subsequent interviews (Table 1). Patients were asked whether they had experienced any of the 19 symptoms from the time of symptom onset to the time of the interview. Additionally, the patient’s baseline characteristics were documented, including past medical history, medication use, and social history.

**Table 1.** Proportion of SARS-CoV-2**-**positive patients who experienced specific symptoms and symptom constellations on days 1, 3, 7, and 14 from symptom onset. The cumulative incidence of the specific symptoms by day 14 is shown in the last column. All values are reported as percentages with 95% confidence intervals (CI).

| Constellation of symptoms | Specific symptoms | Proportion of patients experiencing a symptom per day (%; 95%CI) |  |  |  | Cumulative incidence by day 14 (%) |
| --- | --- | --- | --- | --- | --- | --- |
|  |  | Symptom onset | Day 3 | Day 7 | Day 14 |  |
| Generalized |  | 82 (80-85) | 78 (75-80) | 65 (64-67) | 41 (40-42) | 93 (91-94) |
| - | Fever<br>≥38°C | 42 (36-48) | 33 (29-37) | 18 (17-20) | 6.6 (5.7-7.6) | 47 (45-50) |
| - | Rigor, chills | 32 (27-38) | 24 (21-28) | 13 (12-14) | 4.9 (4.3-5.4) | 43 (40-45) |
| - | Headache | 51 (47-56) | 46 (42-49) | 35 (34-37) | 21 (20-22) | 73 (70-75) |
| - | Myalgia | 55 (51-60) | 45 (42-49) | 27 (26-29) | 10 (10-11) | 62 (59-64) |
| - | Lethargy | 38 (34-42) | 38 (34-41) | 36 (35-38) | 27 (26-29) | 74 (72-77) |
| - | Loss of appetite | 24 (20-28) | 23 (20-26) | 20 (19-22) | 11 (11-12) | 46 (43-49) |
| Upper respiratory |  | 63 (59-67) | 60 (57-63) | 54 (53-56) | 37 (36-38) | 87 (85-89) |
| - | Rhinorrhea | 33 (28-38) | 28 (26-31) | 21 (20-22) | 12 (11-13) | 55 (53-58) |
| - | Sore throat | 33 (29-38) | 27 (25-30) | 18 (17-19) | 10 (9-11) | 45 (43-48) |
| - | Dysosmia | 21 (18-25) | 24 (21-26) | 28 (27-29) | 24 (22-25) | 56 (54-59) |
| - | Dysgeusia | 24 (21-28) | 26 (24-29) | 30 (29-32) | 24 (23-25) | 58 (55-60) |
| Lower respiratory |  | 62 (58-65) | 60 (58-63) | 56 (55-58) | 43 (42-45) | 80 (78-82) |
| - | Non-productive cough | 49 (45-53) | 45 (42-48) | 37 (36-39) | 24 (23-26) | 66 (63-68) |
| - | Productive cough | 13 (11-16) | 14 (12-17) | 16 (15-17) | 15 (14-16) | 39 (37-42) |
| - | Any dyspnea | 25 (22-29) | 26 (24-29) | 28 (26-29) | 22 (21-23) | 52 (50-54) |
| - | Dyspnea at rest | 5.0 (3.8-6.7) | 5.0 (4.0-6.2) | 4.8 (4.2-5.5) | 3.7 (3.3-4.2) | 16 (14-18) |
| Gastro-intestinal |  | 30 (26-35) | 27 (25-30) | 22 (20-23) | 11 (10-12) | 50 (47-52) |
| - | Nausea | 13 (10-17) | 12 (10-14) | 10 (9-11) | 4.9 (4.4-5.5) | 26 (24-28) |
| - | Vomiting | 3.3 (1.9-5.4) | 2.6 (1.7-3.9) | 1.6 (1.1-2.2) | 0.7 (0.4-1.0) | 4.9 (3.9-6.1) |
| - | Abdominal pain | 11 (9-14) | 9.3 (7.9-11) | 6.4 (5.7-7.1) | 3.7 (3.3-4.2) | 24 (22-26) |
| - | Diarrhea | 14 (11-18) | 13 (11-15) | 10 (9.3-11) | 6.0 (5.5-6.6) | 29 (27-32) |

Based on the symptoms documented during the interviews, patients were classified into one of three categories of clinical severity: low severity, defined as mild and improving symptoms; moderate severity, defined as mild dyspnea, cough or fever for less than five days; and high severity, defined as worsening dyspnea, worsening cough, high or persistent fever for five days or longer, or severe fatigue. The frequency of follow-up interviews ranged from daily to every fourth day, depending on clinical severity, age, and underlying conditions. Patients with concerning symptoms were referred for evaluation at the COVID-19 Clinic.

Patients were discharged from telehealth monitoring when they met both of the following criteria: 14 days had passed since the diagnosis of COVID-19, and they had been asymptomatic for the past seven consecutive days.

### Case definitions

Patients were considered symptomatic due to COVID-19 using three different definitions: 1) Reporting any of the 19 symptoms; 2) WHO case definition of suspected COVID-19, which included fever and at least one other symptom of acute respiratory infection, e.g. cough or shortness of breath^19^; 3) Centers for Disease Control and Prevention (CDC) interim case definition of COVID-19, which included either two of the following symptoms: fever, rigor, myalgia, headache, sore throat, dysosmia or dysgeusia, or one of the following symptoms: cough, shortness of breath or difficulty breathing.^20^ Patients who did not fulfill these definitions at the time of diagnosis or during follow-up, were determined to be asymptomatic. Otherwise, they were considered presymptomatic at the time of diagnosis.

### Statistical analysis

The incidence of COVID-19 was calculated by age and sex, using both the Icelandic population and all SARS-CoV-2 tested individuals as denominators. Patients were followed until discharge from the telehealth monitoring service, hospital admission, death or end of the study period (May 22, 2020). Patients enrolled prior to the implementation of the standardized data entry form (March 17, 2020) were excluded from the analysis of symptoms and symptom progression.

The progression of COVID-19 symptoms was analyzed using parametric cure-mixture models and logistic regression. The time frame used in the analyses was days from symptom onset to discharge from telehealth monitoring. The data were interval censored due to the intermittent schedule of the interviews. To account for this, the cumulative incidence of symptom occurrence was estimated with the non-parametric Turnbull estimator and a parametric cure-mixture model using the log-logistic distribution. The proportion of patients experiencing a specific symptom per day was estimated employing logistic regression that allowed for non-linear effects using a three-knot restricted cubic spline. Missing information between interviews was addressed by multiple imputation using chained equations, performed by additive regression, bootstrapping and predictive mean matching procedure.^21^ The number of imputations equaled the highest proportion of missing data for any variable, multiplied by 100. The added uncertainty due to imputation was fully accounted for in the logistic regression models. The result was compared to a complete-case analysis and naive up-down filling procedure.

All statistical analyses were performed in R version 3.6.3.

## Results

### Demographics and clinical characteristics

A total of 45,105 individuals (12% of the Icelandic population) underwent 47,800 tests for SARS-CoV-2 from January 31 to April 30, 2020. Of those, 18,023 (37.7%) were carried out as part of targeted testing and 29,785 (62.3%) were done as part of population screening. Altogether, 1797 patients experienced 1911 positive tests. The population incidence of COVID-19 was 4.9 cases per 1000 individuals, and 40.0 per 1000 tests performed were positive. This rate differed between targeted testing (1703 of 18,023 samples positive or 94.4 per 1000 tests performed) and population screening (208 of 29,785 samples positive or 7.0 per 1000 tests performed). Among tested individuals, the proportion of females (58%) was higher than in the general population (49%). However, the sex distribution was equal among patients who tested positive (Supplemental Figure 1).

All patients who tested positive were enrolled in telehealth monitoring or admitted to hospital, with no exceptions. The clinical progression from symptom onset for the 1797 positive individuals is shown in Figure 1. Thirty-two of those patients were diagnosed after being admitted to hospital of whom one was diagnosed post-mortem (Figure 2). In total, 101 (5.7%) patients were hospitalized, 27 (1.5%) were admitted to intensive care and 16 (0.9%) required mechanical ventilation. The median length of hospital stay was 8 days (IQR, 3.5-19) and 10 (0.6%) patients died.

**Figure 1.**
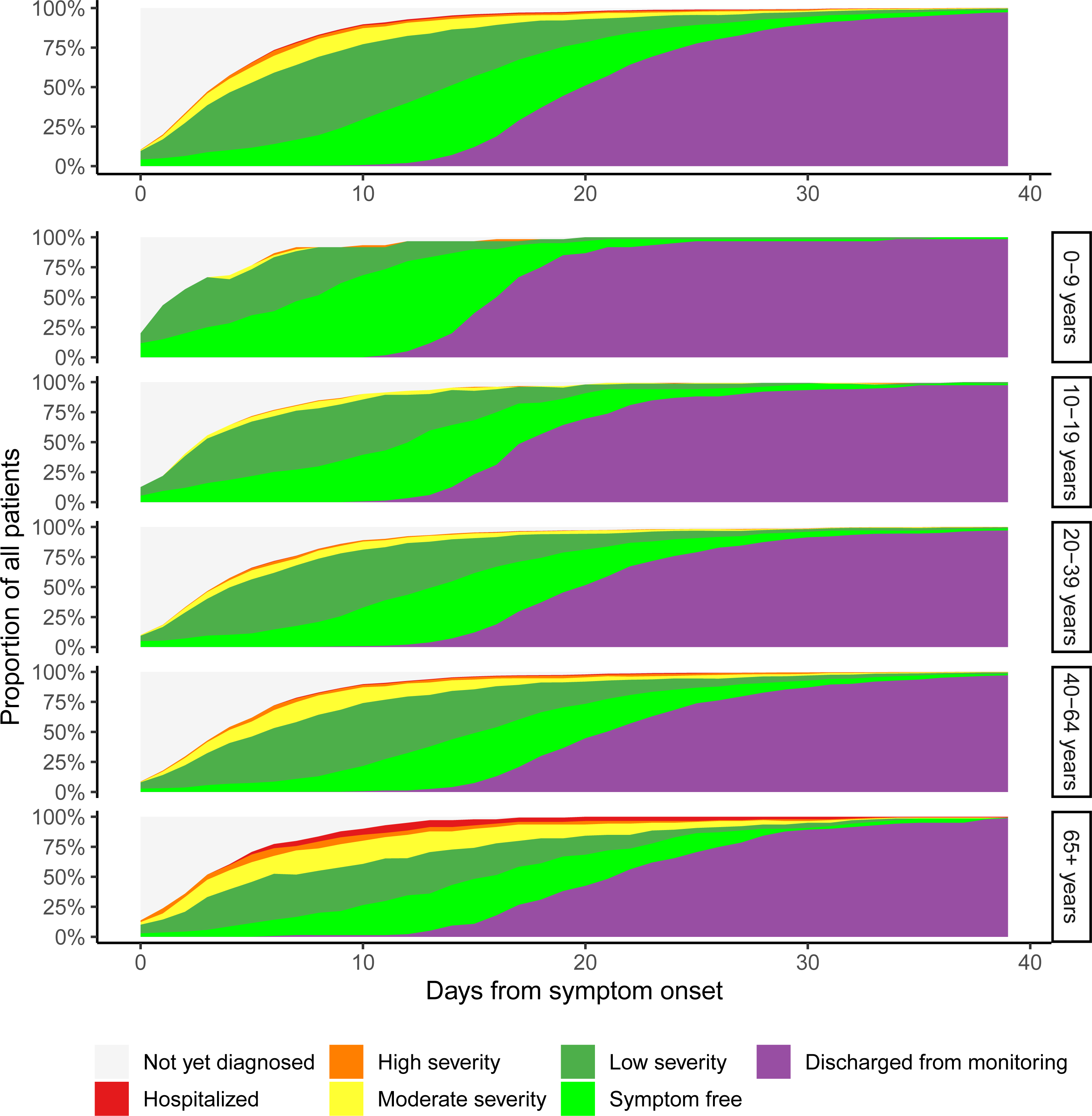
Changes in severity of symptoms from onset to end of follow-up among patients with COVID-19. The category “not yet diagnosed” comprises patients yet to be enrolled in telehealth monitoring, and their clinical severity had therefore not been evaluated. The severity of disease was categorized as low (mild and improving symptoms), moderate (mild dyspnea, cough or fever for less than 5 days), and high (worsening dyspnea, worsening cough, high or persistent fever for 5 days or longer, or severe fatigue). The top panel includes all patients. The other panels show subsets of patients by age group; 0-9, 10-19, 20-39, 40-64, and 65 years of age and older.

**Figure 2.**
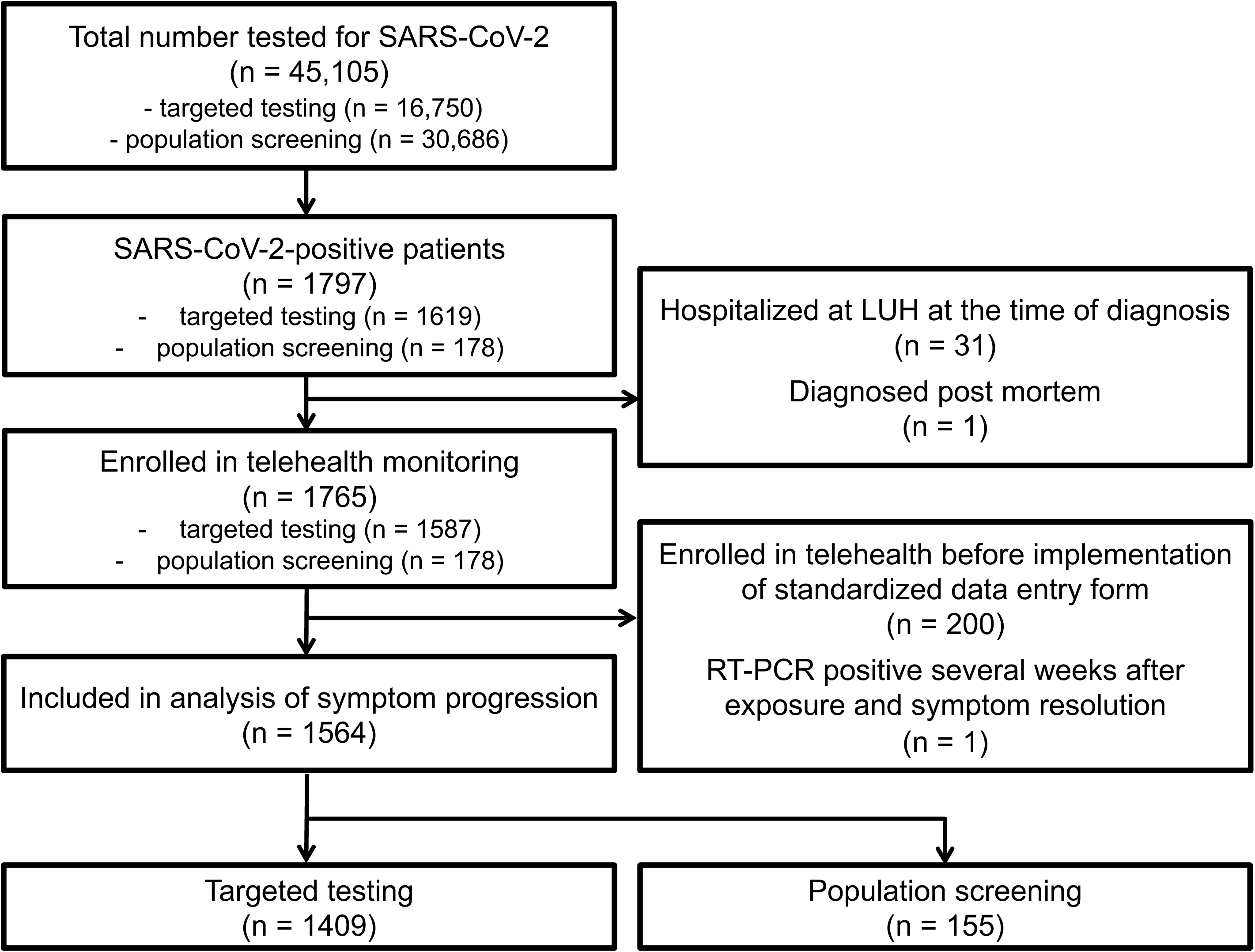
Flow-chart describing the derivation of the study cohort.

### Symptoms at diagnosis

Of the 1797 SARS-CoV-2-positive patients, 41.5% had experienced fever at diagnosis. Other symptoms such as cough (59.2%), dyspnea (30.4%), and gastrointestinal symptoms (32.3%) were also commonly reported. A total of 100 (5.7%) patients reported no symptoms at diagnosis, which was more common among those who were diagnosed through population screening (22.0%) as compared to targeted testing (5.5%). The total number of documented symptoms at diagnosis was lower among patients diagnosed before the implementation of standardized data collection compared to those diagnosed following the implementation (Supplemental Figure 2). Therefore, this latter group was used to study the symptom development and progression in detail (Figure 2).

Of the 1564 (87%) patients who were followed using the standardized data entry form, 791 (51%) were female and the median age was 40 years (IQR, 26-53; range, 0-103). The distribution of age and sex is shown in Supplemental Figure 1. Among these patients, 1055 (67.5%) were classified as having low disease severity throughout their follow-up, 55 (3.5%) were admitted to hospital and 13 (0.8%) required intensive care, six (0.4%) of whom received mechanical ventilation. Two (0.1%) of these patients died. The patients were followed for a median of 15 days (IQR, 14-18) and contributed 69.2 person-years of follow-up time. A total of 1509 patients completed telehealth follow-up from diagnosis until discharge. The observation time of the remaining 55 patients who were hospitalized was censored at the time of admission, after a median follow-up of 4 days (IQR, 2.5-8). Demographic information by subgroup of included and excluded patients is presented in Supplemental Table 1.

Of the 1564 patients with standardized symptom documentation, 42.7% had experienced fever at diagnosis. Cough (60.1%), dyspnea (31.7%), and gastrointestinal symptoms (35.7%) were also commonly reported. Eighty-three (5.3%) patients reported no symptoms at diagnosis, which was more commonly observed among those diagnosed through population screening (24.5%) as compared with patients diagnosed by targeted testing (3.2%). Of the patients who were asymptomatic at diagnosis, 49 (59.0%) remained asymptomatic throughout their telehealth monitoring, while the remaining 34 (41.0%) patients developed symptoms after a median of 3 days (IQR, 3-4.75).

Using the CDC case definition, 216 (13.8%) patients would be categorized as not having symptoms consistent with COVID-19 at the time of diagnosis. This was more common among patients diagnosed through population screening (40.6%) than by targeted testing (10.9%). Seventy-two (4.6%) of those patients developed symptoms compatible with the CDC case definition at a median of 5 days (IQR, 3-6) from diagnosis, whereas the other 144 (9.2%) never met the CDC criteria. Similarly, 349 (22.3%) patients did not fulfill the WHO case definition at the time of diagnosis, a finding that was more commonly observed among patients diagnosed through population screening (45.8%) than by targeted testing (19.7%). The WHO criteria were later met by 115 (7.4%) patients at a median of 4 days (IQR, 3-6) from diagnosis, while the remaining 234 (15.0%) patients never fulfilled the criteria.

The cumulative incidence and proportion of patients meeting CDC and WHO criteria by number of days from symptom onset is shown in Supplemental figure 3. Among the 216 individuals who did not meet CDC criteria at diagnosis, four (1.9%) were hospitalized later in the course of their disease, and one required mechanical ventilation. Similarly, four (1.1%) of the 349 individuals who did not fulfill the WHO criteria at diagnosis were admitted to hospital for illness related to COVID-19, two of whom required intensive care.

### Symptom development and progression assessed by multiple imputation

The median time from symptom onset until RT-PCR diagnosis and enrollment interview was four days (IQR, 2-7) and a median of six (IQR, 4-8) interviews were conducted per patient during telehealth monitoring. The median time between interviews was two days (IQR, 1-3). No data were missing for the days during which interviews occurred. Symptoms occuring on days during which interviews were not conducted were imputed, and the imputation procedure was repeated 92 times. The proportion of patients with missing information on symptoms was highest during the first days following symptom onset (Supplemental Table 2). The proportion of patients experiencing specific symptoms per day from symptom onset was calculated using multiple imputation logistic regression, which produced acceptable results as compared with complete-case analysis and a naïve up-down filling procedure (Supplemental Figures 4-7).

### Symptoms at disease onset

As shown in Table 1, the most common symptoms at the onset of COVID-19 were myalgia, headache, and non-productive cough, observed in 51% (95%CI, 47%-55%), 49% (95%CI, 45%-53%), and 55% (95%CI, 51%-60%), respectively. However, 82% (95%CI, 80%-85%) of patients experienced at least one generalized symptom and 63% (95%CI, 59%-67%) at least one upper respiratory symptom. Compared with patients diagnosed by targeted testing, those diagnosed through population screening were less likely to have experienced cardinal symptoms of COVID-19 at disease onset, including fever (30% vs. 43%), cough (47% vs. 59%), dyspnea (18% vs. 26%) and gastrointestinal symptoms (23% vs. 31%). Other symptoms according to testing protocol are shown in Supplemental Figures 8-11.

The proportion of patients experiencing specific symptoms by sex and age group is displayed in Supplemental Figures 12-15. The initial presentation of COVID-19 varied only slightly between the sexes. The proportion experiencing fever or gastrointestinal symptoms at onset was similar between age groups, but cough and dyspnea were more common among older individuals (Supplemental Figure 14).

Differences in symptoms at onset between hospitalized and non-hospitalized patients are shown in Supplementary Figures 16-19. Several symptoms at disease onset were more common among patients who were later admitted to hospital, including generalized symptoms (94% vs 82%), lower respiratory symptoms (71% vs 62%) and gastrointestinal symptoms (48% vs 29%). However, the proportion of patients who experienced upper respiratory symptoms was lower, 48% compared with 64% of those who were never hospitalized.

### Progression of symptoms

By day 21 from disease onset, the most commonly experienced symptoms were lethargy, headache, and productive or non-productive cough, noted in 74% (95%CI, 72%-77%), 73% (95%CI, 70%-75%) 73% (95%CI 70%-75%), respectively. Overall, 93% (95%CI, 91%-94%) had experienced at least one generalized symptom, 87% (95%CI, 85%-89%) at least one upper respiratory symptom, and 80% (95%CI, 78%-82%) at least one lower respiratory symptom. The cumulative incidence of fever, dyspnea, and gastrointestinal symptoms were 49%, 52%, and 53%, respectively (Figure 3). The proportion of patients experiencing each symptom by days from symptom onset is shown in Figure 4. Of the 741 patients who experienced fever at any time during the course of the disease, 630 (85%) had done so by day 3. Furthermore, 902 of 1140 patients (79%) had developed cough by day 3, 427 of 826 (52%) had developed dyspnea and 485 of 788 (62%) had developed any gastrointestinal symptom by day 3 (Figure 3). These proportions were comparable for both sexes and all age groups (Supplemental Figures 20-31).

**Figure 3.**
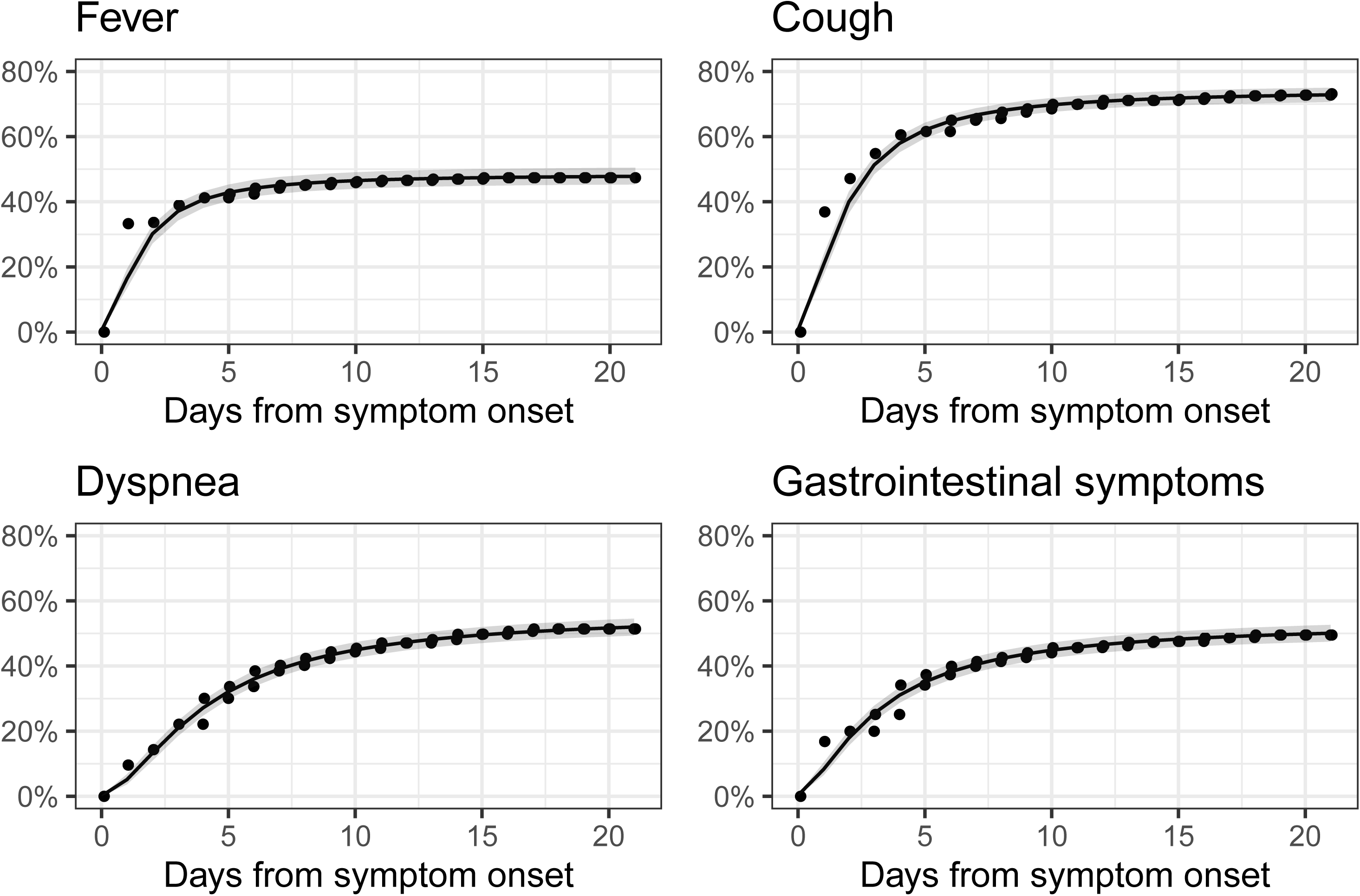
Cumulative incidence of specific symptoms experienced by SARS-CoV-2-positive patients by days from symptom onset. The non-parametric Turnbull estimate of the cumulative incidence is depicted in black points and the parametric cure-mixture estimate is illustrated with a black line with the 95% confidence interval shown as a shaded grey area.

**Figure 4.**
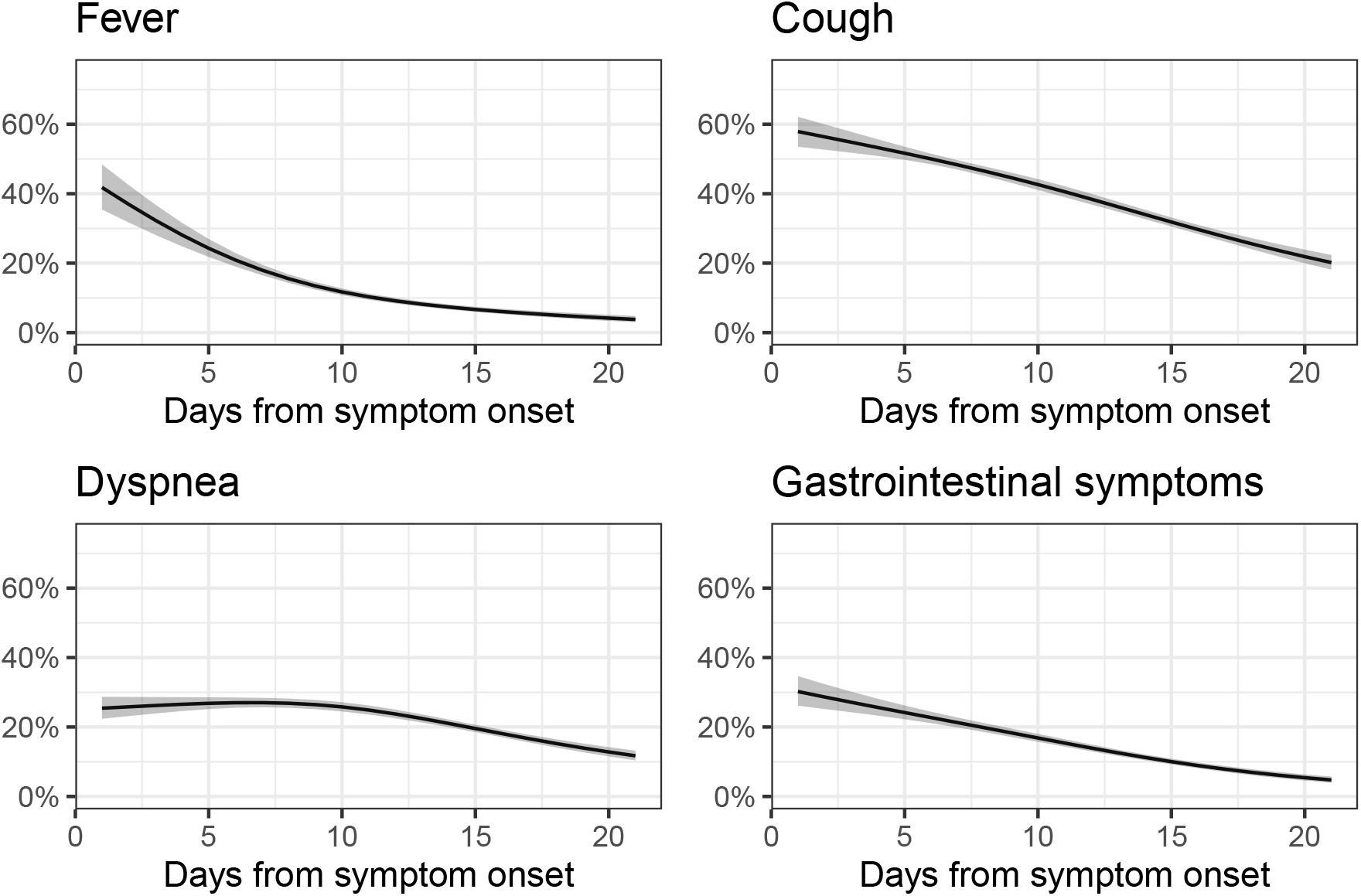
Proportion of SARS-CoV-2**-**positive patients who experienced specific symptoms by days from symptom onset. The logistic regression estimate of the proportion is illustrated with a black line and the 95% confidence interval shown as a shaded grey area.

Of the 19 symptoms, only dysosmia and dysgeusia were more common later in the disease course than at symptom onset. Both symptoms peaked on day eight from diagnosis. The trend was most pronounced among patients aged 25 to 55 years and was more marked among females (Supplemental Figure 13). Other symptoms attributed to COVID-19 were most prevalent during the onset of the disease. No symptom exhibited a bimodal pattern (Supplemental Figures 12-15). The cumulative incidence of each specific symptom was lower among patients diagnosed through population screening compared to targeted testing, except for rhinorrhea and vomiting (Supplemental Figures 32-35). By day 21, a large proportion of patients who were ever admitted to hospital for COVID-19 had experienced fever, dyspnea and and gastrointestinal symptoms, or 91% (95%CI, 71%-95%), 83% (95%CI, 64%-91%), and 84% (95%CI, 66%-91%), respectively (Supplemental Figures 36-39).

## Discussion

In this study we present the clinical characteristics of COVID-19 in a national population-based cohort. Due to aggressive contact tracing and widespread virological testing it is likely that the cohort includes the majority of symptomatic cases in the population. This assumption is supported by the low prevalence of the disease detected by random population screening (0.6%).^15^ Our prospectively collected data regarding symptoms and disease progression among patients who tested positive for SARS-CoV2 in Iceland revealed that 49% of patients experienced fever, 73% cough, and 52% dyspnea. At the time of diagnosis, 5% were completely asymptomatic, 13% did not meet the CDC case definition, and 22% did not fulfill the WHO criteria.

The comprehensive, nationwide characterization of COVID-19 symptoms was facilitated by broad access to diagnostic testing in Iceland. The Icelandic healthcare system is a single-payer system with a universal government-run health insurance provider. The SARS-CoV-2 RT-PCR test was free of charge for both targeted testing and population screening, resulting in over 12% of the population tested, which was higher than in any other country during the study period.^16^ As a result, we were able to describe the true spectrum of COVID-19, while previous studies were largely based on hospitalized cohorts or cases identified in the setting of more restrictive testing.^6,12,22^ Furthermore, subgroup analysis allowed us to quantify the degree by which cohorts with only hospitalized patients might overestimate the presence of specific symptoms, for instance fever, which was considerably more common at symptom onset in patients who were later admitted to hospital (74%) than in patients who never were hospitalized (40%).

The proportion of patients with COVID-19 who were completely asymptomatic has been a focus of interest during the pandemic with implications for the risk of disease dissemination. A recent report from Iceland examining an overlapping cohort suggested that 43% of patients were asymptomatic at the time of sampling.^15^ This estimate was derived from 43 out of 100 RT-PCR positive cases reporting no symptoms at the time of diagnosis among 13,080 participants in a population screening program, which included both open-invitation and random invitation screening. Follow-up information on the patients’ symptoms was not available to the authors^15^, possibly resulting in an overestimation of asymptomatic individuals. In the current study, 83 patients reported no symptoms at the time of diagnosis, approximately half of whom developed symptoms in the ensuing days. Thus only 3.1% of diagnosed cases remained completely symptom-free during follow-up. However, as some degree of suspicion of COVID-19 was needed to prompt an individual to be tested, symptomatic patients are likely to be overrepresented in our sample. Estimating the true proportion of SARS-CoV-2-positive patients who never develop symptoms is difficult, as asymptomatic patients are unlikely to be diagnosed outside of random population screening, and differentiating pre-symptomatic from asymptomatic patients requires longitudinal follow-up. Gudbjartsson et al. found that 13 out of 2283 randomly sampled Icelanders were SARS-CoV-2positive, of which 7 (53.8%) reported no symptoms at the time of diagnosis. We have shown that 59.0% of patients who are symptom free at diagnosis, never develop symptoms. Based on these observations, the rough estimate of the proportion of truly asymptomatic SARS-CoV-2-positive Icelanders is approximately 30%.

Of symptomatic patients, most experienced only minor symptoms. Only 22% of patients developed moderate symptoms, 8% severe symptoms, and 3.5% were hospitalized. The standardized prospective recording of clinical symptoms made it possible to evaluate the sensitivity of the widely used CDC^20^ and WHO^19^ case definitions for the diagnosis of COVID-19 throughout the course of the disease. By applying these definitions, we demonstrate that a substantial number of cases would have been missed; approximately 9% by the CDC criteria and 15% by the WHO criteria. The identification of additional 4% and 7% of cases would have been delayed by a median of 5 and 4 days, respectively. These are concerning results with immediate implications for current efforts to curtail the pandemic. Our data show that most patients have mild symptoms that may not have prompted the consideration of COVID-19 by either patients or health care providers in more resource-limited settings, and indicates a need for revising and widening the CDC and WHO case definitions to increase their sensitivity.

Symptoms observed among patients with mild forms of COVID-19 have previously been examined in a multicenter European study of 1420 RT-PCR positive patients who answered a questionnaire.^10^ Severely ill patients were excluded and the remaining cohort was predominantly female (68%), young (94% were <60 years of age), and biased towards healthcare workers (31% of the group).^10^ While these results are not easily generalizable to the entire population, the investigators found that only 7% of patients required hospitalization^10^ compared to 3.5% in the present study. Headache, loss of smell, and nasal obstruction were the most common symptoms identified.^10^ Although these symptoms were also frequently identified in our cohort, we found cough and myalgias to be more common. The predominance of loss of smell identified in the aforementioned study agrees with our observation that olfactory symptoms are most common in younger age groups.^10^

We found that slightly less than 50% of patients developed fever during the course of the disease, already present in 85% of those by day 3. This is consistent with the study by Lechien et al. in which fever ≥38.0°C was reported in 45.4% of cases^10^, while it is higher than was reported by Guan et al. (21.7%)^5^ and Goyal et al. (25%)^12^. A meta-analysis by Sun et al. found that 89% of COVID-19 patients had a fever ≥37.3°C^6^, but this definition of fever is rarely used in clinical practice. Over the follow-up period, 70% of patients experienced cough which is consistent with the findings of Lechien et al. who observed cough in 63% of cases^10^. In total, 52% of patients reported any dyspnea and only 13% reported dyspnea at rest during the disease. The reported incidence of dyspnea ranges from 22-49%^7,10,23^ but most previous studies do not differentiate between dyspnea at rest and on exertion. Gastrointestinal symptoms were common, reported by almost half of patients at some point during the first 14 days. Abdominal pain (22%) and diarrhea (28%) were frequent as in previous studies.^12,24^ Interestingly, although one-fourth of patients experienced nausea, vomiting was rare.

Our findings indicate a lower rate of hospital admissions and mortality in Iceland compared with many other countries. The reasons for these disparate outcomes are likely multifactorial. Iceland has a relatively young population, with 85.8% younger than 65 years, compared with 77.1% in Italy, 80.6% in Spain, 81.6% in the United Kingdom, and 83.5% in the USA.^25,26^ This, in addition to a strong emphasis on limiting exposure of elderly and multimorbid individuals, resulted in a low median age of confirmed COVID19 cases of 40 years (IQR, 26-53) in Iceland compared to 51 years (IQR, 36-65) among all cases reported to the WHO^27^ and 48 years (IQR, 33-63) in the USA.^28^ Different rates of other recognized risk factors for worse outcomes in COVID-19 are unlikely to explain this difference as they have a similar distribution in Iceland as in other countries, for instance 28.8% of Icelandic adults have hypertension^29^ and 27% are obese.^30^

This study does have some limitations. In order to accurately describe symptom progression, cases diagnosed before the implementation of the standardized clinical data entry form were excluded from the analysis of symptom development, representing 11% of all SARS-CoV-2-positive cases in Iceland. The date of implementation of the standardized data entry form was not influenced by the clinical characteristics of the patients being diagnosed, and therefore should not introduce bias. Furthermore, the demographics and clinical characteristics of excluded cases were largely comparable to those that were included in the study. Another limitation is that daily standardized documentation of symptoms was not available during hospital admission. This could conceivably lead to an underestimation of severe symptoms such as dyspnea. However, only 3.5% of the included patients were hospitalized, and symptoms prior to hospitalization were included in the analysis. It is important to note that the data were based on self-reported symptoms via telephone calls. This shortcoming is mitigated by the fact that experienced nurses and physicians conducted the interviews. A principal strength of the study is its population-based approach, which included all confirmed cases in the country during the study period, regardless of their need for medical care.

### Conclusion

This study describes the symptomatology and clinical severity of the initial phase of COVID-19 in Iceland. The incidence of COVID-19 was high due to extensive testing of both symptomatic and asymptomatic individuals, while disease severity was lower than previously reported. Symptoms such as fever and dyspnea were less frequent than has been observed in earlier studies. Our findings suggest that both the CDC and WHO case definitions of COVID-19 lack sensitivity and miss a substantial proportion of patients, including cases who later develop severe disease.

## Data Availability

The data that support the findings of this study are available on reasonable request from the corresponding author. The data are not publicly available as they contain information that could compromise the privacy of research participants.

## Authors’ contributions

Elias Eythorsson - Conception and design, acquisition of data, analysis and interpretation of data and drafting of the manuscript.

Dadi Helgason - Conception and design, acquisition of data, interpretation of data and drafting of the manuscript

Ragnar Freyr Ingvarsson - Conception and design, and drafting of the manuscript

Helgi K Bjornsson - Conception and design, acquisition of data and drafting of the manuscript

Lovisa Bjork Olafsdottir - Conception and design, acquisition of data and drafting of the manuscript

Valgerdur Bjarnadottir - Acquisition of data and revision of manuscript

Hrafnhildur Linnet Runolfsdottir - Acquisition of data and revision of manuscript

Solveig Bjarnadottir - Acquisition of data and revision of manuscript

Arnar Snaer Agustsson - Acquisition of data and revision of manuscript

Kristin Oskarsdottir - Acquisition of data and revision of manuscript

Hrafn Hliddal Thorvaldsson - Acquisition of data and revision of manuscript

Gudrun Kristjansdottir - Acquisition of data and revision of manuscript

Brynja Armannsdottir - Conception and design and revision of manuscript

Agnar Bjarnason - Conception and design and drafting of the manuscript

Birgir Johannsson - Conception and design and revision of manuscript

Olafur Gudlaugsson - Conception and design and revision of manuscript

Magnus Gottfredsson - Conception and design and revision of manuscript

Martin I Sigurdsson - Conception and design, interpretation of data and revision of manuscript

Olafur S Indridason - Conception and design, interpretation of data and revision of manuscript

Runolfur Palsson - Conception and design, interpretation of data and revision of manuscript

All authors gave final approval of the submitted manuscript

